# OpenPFx: Evaluating the Ability of LLMs to Create Patient-Friendly Explanations of Radiological Incidental Findings

**DOI:** 10.1101/2025.10.10.25337597

**Authors:** Joseph Hentel, Kurt Teichman, George Shih

## Abstract

The 21st Century Cures Act mandates patient access to electronic health information, yet radiology reports often remain inaccessible due to specialized terminology and widespread low health literacy. This study evaluates large language model (LLM)–based workflows for generating patient-friendly explanations (PFx) of incidental MRI findings. Four approaches—zero-shot, few-shot, multiple few-shot, and agentic—were benchmarked using ICD-10 code alignment for accuracy and Flesch Reading Ease scores for readability. Across 407 outputs per workflow, the agentic method demonstrated the strongest overall performance, achieving a sixth-grade reading level and the highest accuracy. Compared with prior work limited by small sample sizes or suboptimal readability, these results indicate that structured, agent-based LLM workflows can improve both clarity and diagnostic consistency at scale. By translating complex radiology findings into accessible language, AI-generated PFx provide a scalable strategy to reduce health literacy disparities and advance the Cures Act’s goal of making medical data both transparent and usable for patients.

## 1 Introduction

The 21st Century Cures Act grants patients access to all electronic medical data, including medical reports [1]. Since its implementation, there has been a drastic increase of the number of patients accessing Electronic Health Reports (EHR) [2]. While increased access to medical reports is meant to empower the patient, accessed reports are mainly written for referring physicians, leaving patients unable to understand them [3]. Data suggests only 12% of American adults have “proficient” health literacy [4]. Low health literacy has been repeatedly recognized as a stronger predictor of poor health than age, income, education-level, or race [5]. Affected patients have been found to misread medication labels, be unable to make informed medical decisions, and have higher emergency department (ED) readmission rates [6]. One study concluded 88% of readmissions within a 30-day period were due to a misunderstanding in their discharge summary [7]. Problems that arise from patient illiteracy strain the healthcare system, causing an estimated additional 238 billion dollars per year in the United States alone [8]. Low health literacy is not random; instead, it highlights existing systemic inequities. An estimated 65% of Hispanic adults and 58% of African-American adults have a basic or below-basic level, compared to just 28% of White adults [9]. Literacy disparities are not only racial. A 2019 study quantified that education and income account for approximately 37% and 30%, respectively, of total disparities in health literacy across American adults [9]. As a result, underserved communities with low health literacy may be at a significant disadvantage.

Addressing systemic inequities requires solutions that not only consider medical complexity, but also the accessibility of information. Patient-friendly medical reports aim to explain complex medical information in a simplified manner while maintaining accuracy [10]. Adapting medical reports to patients’ reading levels could minimize the detrimental effects of poor health literacy. Patient-friendly medical reports can also make doctors’ use of time more efficient. Appointments with the sole purpose of explaining medical terminology would be significantly reduced [11]. Currently, patient-friendly medical reports must be generated by a medical professional. Despite their proven benefits, writing them is too time consuming to complete for every patient [12]. Due to advancements in generative AI, researchers have experimented with large language models (LLMs) — a type of artificial intelligence that process and generate natural language — to generate patient friendly medical reports [13]. This would automate the process, making patient-friendly medical reports possible [14]. When LLMs were used to generate discharge summaries, the AI-generated summaries hit a 6th grade reading level, meeting the NIH’s recommendation for patient-friendly material [3]. Despite the progress made in readability, the accuracy of these reports is a concern. The generated discharge summaries only had 54% accuracy with 18% being flagged for safety concerns [3]. Currently, most research utilizes zero-shot prompting [15]. In zero-shot prompting, no context such as a template or an example letter is provided, requiring the LLM to rely solely on its previous knowledge [16]. For example, a zero-shot prompt for generating a patient-friendly medical report would consist of the original medical report and a straight-forward prompt to generate a patient friendly explanation. Zero-shot prompting commonly produces reports that necessitate manual refinement prior to being delivered to patients [15].

**Figure 1:**
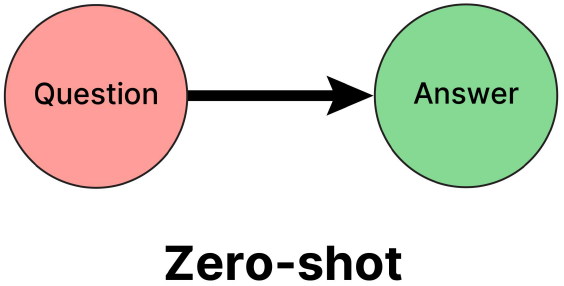
Zero-shot Flowchart

[17] Few-shot prompting provides context such as sample responses or templates, leading to more accurate and tailored outputs [16]. In this context, a few-shot prompt would contain sample medical reports and patient friendly versions of them along with a command to generate the patient-friendly report. While few-shot prompting has the potential to outperform zero-shot, manual refinement of the outputs may still be needed [15].

**Figure 2:**
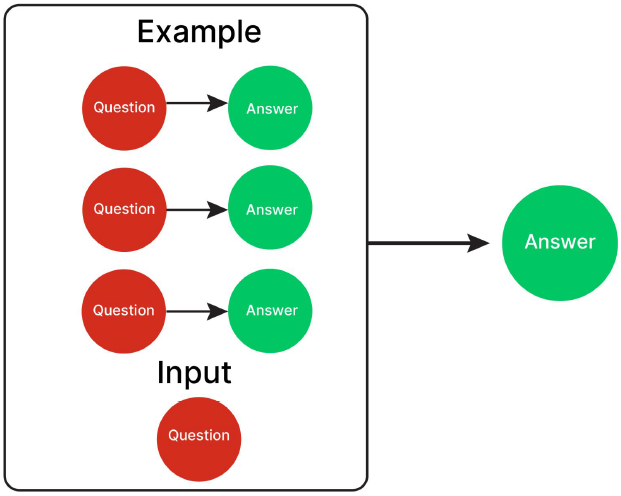
Few-shot Flowchart

[18] Although zero and few-shot prompting serve as important foundations, emerging research indicates that agentic prompting workflows may define the next phase of AI development [19]. An agentic workflow is a structured process that enables AI models, acting as “agents,” to iteratively refine their outputs based on feedback and contextual adjustments [19]. Each agent is a specialized AI system with access to tools designed to perform specific tasks, such as analyzing data, generating outputs, or providing feedback to other agents [19]. Rather than producing a single output in response to a prompt, the workflow involves multiple stages where these agents evaluate and improve their performance collaboratively [19]. This iterative refinement seeks higher accuracy and relevance by addressing errors or ambiguities in earlier outputs [19]. A study found that agentic workflows reduced the need for edits from 68.75% in zero-shot outputs to 18.75% [15]. Agentic AI is rapidly evolving. On top of limited research using agentic AI to translate medical reports, this research is already outdated.

**Figure 3:**
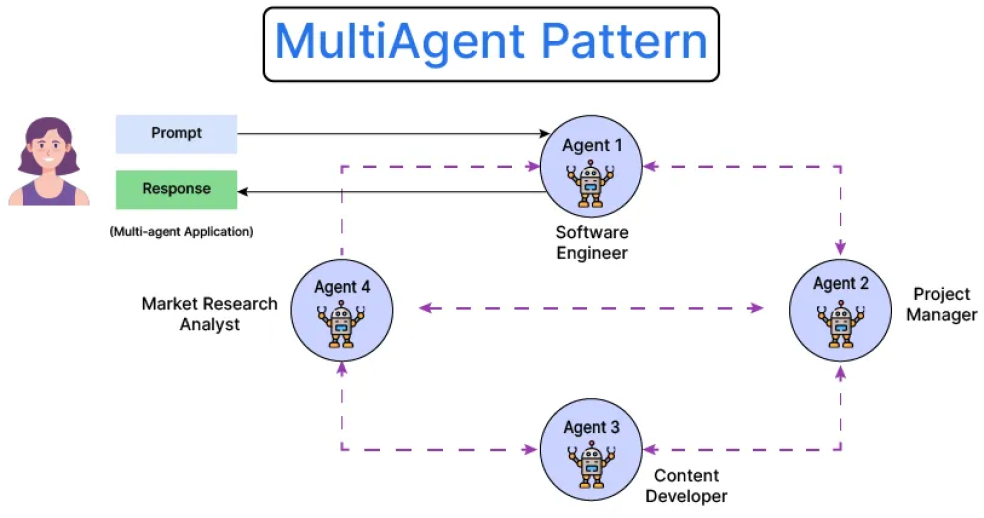
Multi-agent Flowchart

[20] Although AI-generated patient-friendly medical reports have shown promise in improving patient understanding, the challenge becomes even more pronounced when dealing with incidental findings. Incidental findings are unexpected abnormalities discovered in medical imaging that are not related to the reason the imaging was performed [21]. These findings, often unexpected and filled with technical terms, can leave patients confused and anxious about their significance [21]. Potentially even worse, some patients may disregard the finding as it is not related to their original scan and do not seek the necessary treatment [22]. A study found only 17% of ED patients provided with incidental findings scheduled follow-ups, along with 62% in outpatient centers. [22]. Translating these findings into clear, actionable language poses unique difficulties, as they require both medical accuracy and a compassionate tone to relieve anxiety. There is extremely limited research on the ability to simplify incidental findings. In an abstract created prior to conducting this study, few-shot prompting achieved an 8th-9th grade readability, showing promise, but ultimately failing to meet the 6th grade recommendation [23]. These limitations emphasize the need for further research to develop workflows that ensure proper explanations of incidental findings. Identifying the best methods will be essential to improve patient understanding and reduce anxiety.

## 2 Methods

### 2.1 Data Creation

The list of incidental findings was created using ChatGPT 4o. Several research articles detailing the most common incidental findings in MRI were fed to ChatGPT 4o. ChatGPT 4o was then asked to list the incidental findings along with their ICD-10 codes. Codes were manually verified.

### 2.2 LLM Workflows

Four workflows were evaluated: zero-shot, few-shot, multiple few-shot and agentic.

All prompts, including agent system messages, were created using the LangChain Prompt Template to provide a clear, organized format to the prompts. Structured outputs were created using the Pydantic framework.

The zero-shot instruction contained a prompt and other specific directions. It requested for a patient-friendly explanation of a specified incidental finding at a 6th grade reading level. After writing the PFx, the model was told to assign an ICD-10 code to its own explanation. The few-shot instruction was the same as the zero-shot, except it included sample PFx. Multiple few-shot instructed the LLM to generate the response 5 times in a row. This was to test if the LLM would improve its output while generating a response multiple times. The best out of the five responses was selected as the response for multiple few-shot. Selection was determined by an overall score metric where readability was weighed 20% and accuracy 80%.

The agentic workflow was created using the AG2 library. AG2, a community-driven fork of the original Autogen, is an open-source framework for building AI-powered agents. The RoundRobin pattern was selected due to its collaborative nature. The pattern enables agents to communicate freely with one another. To maintain order in the conversation, agents were provided specific handoff instructions with what agents they were permitted to speak to and when.

The agent-based workflow contained four agents: a writer, code labeler, doctor, and readability checker. The writer received the same zero-shot prompt used earlier. After generating the PFx, the writer would send the PFx to the ICD-10 code labeler. The labeler would read the response and assign it an ICD-10 code. Next, a doctor agent would proofread the response for medical accuracy. If accurate, the doctor would send the PFx to the readability checker to ensure a 6th grade readability. If readable, the conversation would end. If the doctor or readability checker deemed the response inadequete, they’d send the response back to the writer with suggestions for revision. Once the conversation ended — either achieving accuracy and readability or hitting the 20-turn limit — the last response outputted by the ICD-10 code labeler would be extracted to be used as the PFx.

**Figure 4:**
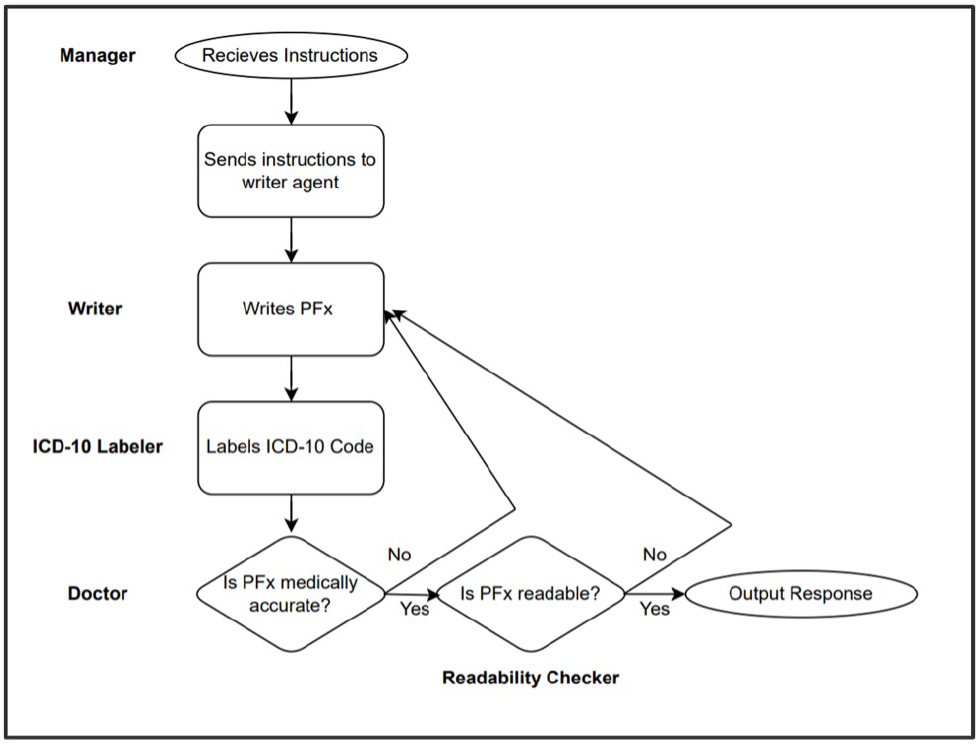
Original Agentic Workflow Flowchart

### 2.3 Evaluation Metrics

Accuracy was measured using ICD-10 code checks. ICD-10 (International Classification of Diseases, Tenth Revision) is a standardized system for classifying and coding medical diagnoses [24]. ICD-10 codes were assigned to the list of incidental findings by ChatGPT 4o, but each code was manually verified to ensure accuracy. Two checks would occur throughout the process. The first check compares the actual ICD-10 code to the code generated during the creation of the PFx. For all three workflows, after the PFx was generated, a separate LLM would read it and assign an ICD-10 code to it. The second check compares the actual ICD-10 code to this code. The two checks were weighted evenly and averaged to get the accuracy of a PFx.

The primary purpose of ICD-10 codes is billing. This requires codes to be highly precise. On top of the general finding, ICD-10 codes specify details such as location, cause, severity, and encounter type. The primary purpose of the explanations generated in this study is to provide the patient with general knowledge, not an ultra-specific details. The first three digits of an ICD-10 code capture the broad diagnostic category. Therefore, only the first three characters could be used to identify the general condition, without penalizing it for missing billing-level specificity that falls outside the project’s scope. A relaxed accuracy metric that only compares the first three characters was created to counter these small differences. This metric prioritizes the general condition over specificity.

Readability was measured using a Flesch Reading Ease Score (FRES). The FRES assigns a score (0-100) based on average sentence length and average word syllable count, with higher scores indicating easier readability [25]. Certain ranges in the scale correspond to a specific grade’s reading level. A score of 80-90 is a 6th grade reading level, so the target FRES score was an 85.[26]

**Figure 5:**
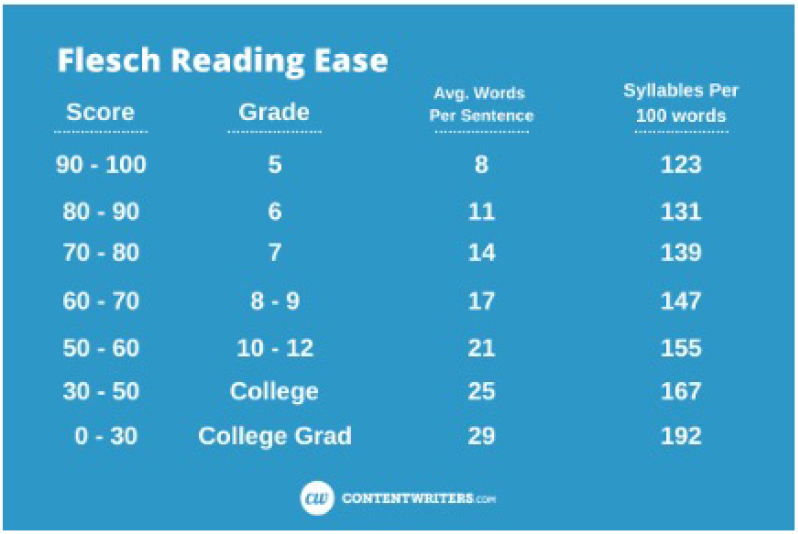
FRES Score Chart

## 3 Results

The agentic workflow outperformed all other workflows in every category.

Word counts were similar in all workflows although the agentic workflow had the highest average (92.12 +-7.81; p < 0.01).

The only workflow to achieve an average of the NIH recommended 6th grade reading level was agentic (84.12 +-7.13; p < 0.01). Zero-shot’s average FRES was just outside the 80-90 range (79.22 +-8.17; p < 0.01) while both few-shot and multiple few-shot were further (65.15 +-9.23 and 65.78 +-8.51, respectively; p < 0.01).

Multiple few-shot did not demonstrate an advantage over generic few-shot. In fact, readability rose an entire grade level (few-shot 65.15 ± 9.23; multiple few-shot 56.32 ± 8.25). For word count, accuracy, and relaxed accuracy, measured results were within 4% of each other. In strict accuracy, few-shot outperformed multiple few-shot (0.58 +-0.47 and 0.56 +-0.47, respectively,; p < 0.01).

In terms of strict accuracy, the agentic strongly outperformed all other workflows (0.70 +-0.36 with next best being few-shot with 0.58 +-0.47; p < 0.01) The advantage was smaller in relaxed accuracy (0.86 +-0.28 and next best few-shot/multiple few-shot with 0.80 +-0.38; p < 0.01).

**Figure 6:**
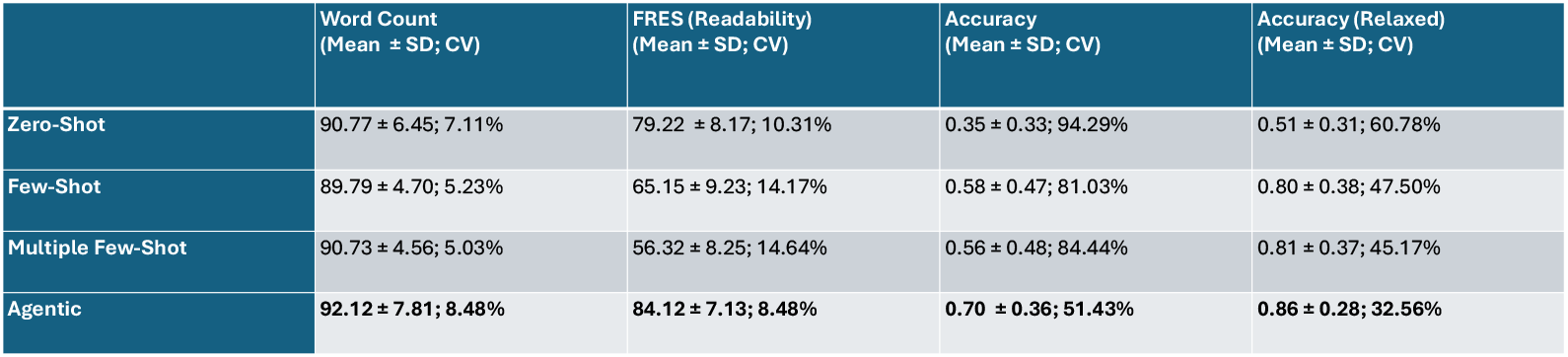
Results Table

**Figure 7:**
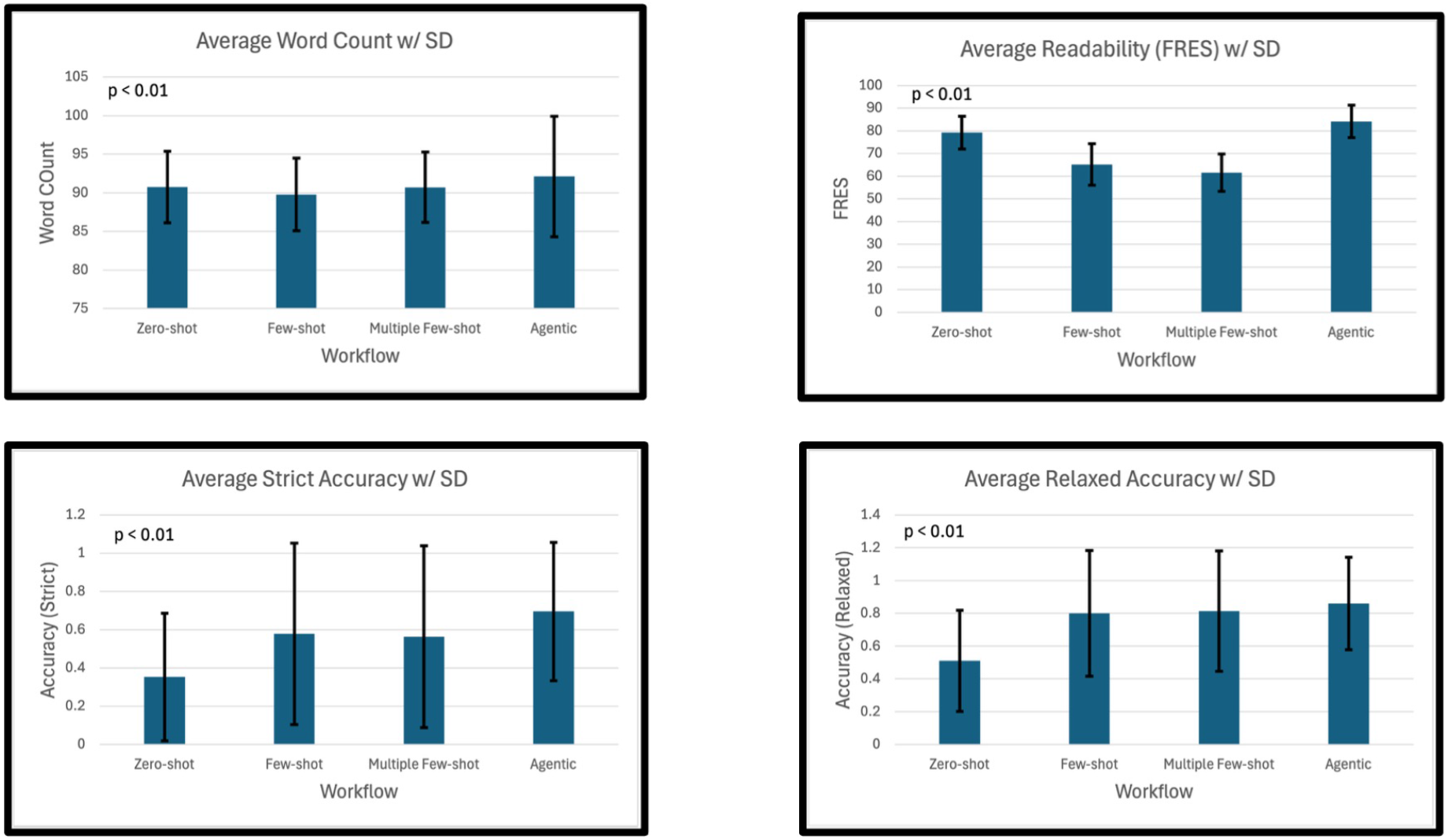
Comparison of Workflows on Different Metrics w/Standard Deviation

## 4 Discussion

### 4.2 Readability

For the agentic workflow, the average FRES was 84.12, putting it right in the middle of the 6th grade range. This shows extremely strong promise for its ability to generate patient-friendly explanations of incidental findings, and overall, explanations of all medical material. Patient-friendly material reading at a 6th grade reading level has the potential to have profound effects on the overall patient experience. A study analyzing the readability of discharge summaries - letters patients receive when discharged from the hospital - determined that approximately 2/3 of patients had a reading level lower than what their summary was written in [7]. This means that 2 out of every 3 people leaving the hospital do not fully understand what is wrong with them. Approximately 54% of the American adult population (ages 16 – 74) read at or below a 6th grade reading level, demonstrating the dire need for the readability of patient material to drop [27]. Although explanations of incidental imaging findings are only a fraction of what a full medical report encompasses, it is a step in the right direction. Improving readability also has implications for health equity. Populations with lower literacy rates — disproportionately affecting racial and socioeconomic minority groups — are the most likely to benefit from explanations written at a sixth-grade level. By closing this readability gap, PFx systems could help reduce disparities in patient understanding and engagement with their care.

### 4.2 Word Count

While the workflows produced similar word counts, the agentic workflow generated the longest explanations on average (92.12 ± 7.81 words). Higher word counts can indicate richer explanations, which may improve patient understanding by providing additional context. However, longer outputs may also risk overwhelming patients with too much information or unnecessary details. The readability results suggest that agentic’s longer explanations did not compromise clarity; in fact, they aligned with the NIH’s recommended sixth-grade level. This suggests that higher word counts, when paired with accessible language, may be beneficial rather than burdensome. Though, outside of the ICD-10 checks, the actual content of the reports was not verified, meaning the reports could contain readable, yet unrelated content.

### 4.3 Accuracy

The accuracy, if measured correctly, still poses a problem with AI-generated explanations. Even with the relaxed accuracy metric, the most accurate workflow, agentic, still only had 84% accuracy. A limitation with measuring accuracy in this study is the reliability of GPT-4o to generate accurate ICD-10 codes. Previous research has found that it is reliable with generating correct codes, but LLMs still have the potential to hallucinate [15]. The relaxed accuracy metric attempted to counteract LLM hallucinations by reducing the number of characters needed to match in the ICD-10 checks. Still, LLMs can provide a completely unrelated code for a response that adequately addresses its specified finding. If the PFx generated by this study were to be implemented in a clinical setting, they should be proofread by a medical professional.

### 4.4 Reproducibility

Although the agentic workflow demonstrated the best average performance across all categories, reproducibility remains a concern. Agentic achieved the lowest variability overall, with a standard deviation of 7.13 in readability (CV ≈ 8.5%), suggesting that it generated consistently understandable explanations. However, accuracy results were much more variable, with CVS of 51% (strict) and 32% (relaxed). This suggests that accuracy is far less reproducible than readability. Part of this variability, however, stems from the structure of the accuracy metric itself. Each explanation was evaluated by only two ICD-10 checks. As a result, accuracy scores could only take on three values: 0, 0.5, or 1.0. This inflates the standard deviation and coefficient of variation, since even small differences in coding led to large changes. The relaxed accuracy metric helped mitigate this effect, reducing the CV compared to strict accuracy (for the agentic workflow, strict accuracy had 51.43% CV, while relaxed accuracy dropped to 32.56%). While this adjustment provided more stable results, the variability remains significantly higher than acceptable for clinical deployment. For clinical applications, reproducibility is critical: patients should not receive markedly different explanations for the same incidental finding. While the agentic workflow reduced variability compared to zero-shot and few-shot methods, its accuracy scores still leave room for improvement. To indicate strong reproducibility, the CV would be below 10%. Future work could address this by incorporating more than two independent code checks per explanation or developing more robust and holistic confidence measures.

### 4.5 Comparative Context

Zaretsky et al. evaluated 50 discharge summaries and found that AI-generated outputs averaged an 6th-grade reading level, well above the NIH’s sixth-grade target, and achieved only 54% accuracy, with 18% of outputs flagged for safety concerns. [3]. Sudarshan et al. reported an agentic workflow that achieved 94.94% ICD-10 accuracy but tested only 16 radiology reports, and their outputs did not consistently meet the sixth-grade readability standard [15]. In contrast, the present study evaluated 407 outputs per workflow and found that while accuracy was lower (70% strict; 86% relaxed), the agentic workflow uniquely achieved an average FRES within the NIH’s sixth-grade target range. Taken together, these findings suggest that while prior studies advanced either accuracy or readability, this project provides stronger evidence of real-world potential by demonstrating both on a much larger scale.

### 4.6 Limitations

On top of previously discussed limitations such as the accuracy and reproducibility, this project has other limitations. First, this none of the generated explanations were reviewed by patients or physicians. This means that there could be content that may not be inaccurate or hard to read, but still unnecessary or misleading. Prior to clinical implementation, reports must be reviewed and approved by medical professionals. Additionally, large language models are rapidly evolving. Although the larger, general use LLMs have are less sensitive to small prompt difference, prompt effectiveness still varies on the type of model. With the release of newer models or even the updating of the same model used, prompts must be retested and refined.

### 4.7 Future Work

Future research should focus on preparing PFx workflows for clinical implementation. A key priority is refining accuracy, both by improving ICD-10 coding reliability and even moving beyond simple code matching to clinically meaningful correctness. This will likely require additional strategies such as fine-tuning, ensemble prompting, or integrating external medical knowledge bases. Clinical deployment would also necessitate robust content verification, where generated explanations are reviewed by physicians or supported with robust confidence scores to prevent unsafe outputs from reaching patients. In addition, research should focus on expanding the scope from isolated incidental findings to full radiology reports. Finally, studies involving patients and clinicians will be essential for testing whether improved readability and accuracy actually translate into better comprehension, trust, and patient outcomes in real-world settings.

## 5 Conclusion

The findings demonstrate that structured, agent-based large language model workflows can generate patient-friendly explanations of incidental findings that are both readable and clinically aligned. Among all workflows tested, the agentic approach most effectively balanced accuracy, clarity, and consistency, producing outputs that met the NIH’s sixth-grade readability recommendation while achieving the highest accuracy. These results underscore the potential of AI-driven systems to operationalize the goals of the 21st Century Cures Act by transforming complex radiology data into information patients can understand and act upon. Broader integration of such workflows may enhance health literacy, promote equity in patient understanding, and advance a more transparent and accessible healthcare system.

## Data Availability

All data produced are available online at: https://github.com/joeyhentel/OpenPFx/

## 6 Acknowledgments

I would like to sincerely thank my mentor Dr. George Shih for introducing me to the topic and providing consultation all throughout the project. I would also like to thank Mr. Kurt Teichman for helping with programming related questions.

## 7.1 OPENPFX WEBSITE

https://openpfx.streamlit.app

## 7.2 GITHUB LINK FOR PROJECT

https://github.com/joeyhentel/OpenPFx

## A ChatGPT 4o PFx Data Generation Prompts

NOTE: Each prompt listed was used in the same conversation in chronological order

NOTE: Citation indicates attached article to prompt

Prompt 1:

[28] [29] [30] [28] [31] [32] [33] [34] [35] [36]

You are a medical researcher tasked with generating a list of incidental findings in MRI. generate 50 MRI incidental findings for head MRIs and their corresponding icd10 codes. make it in a csv format. there should be NO REPEATS. format each row like this. [Body Part, Organ, Finding, ICD-10 Code, ICD-10 Code Description].

Prompt 2-8:

Generate another 50 following the directions I previously provided.

*Prompt 2 was repeated until 400 findings were created*

Matched 400 findings against manually generated list of 25 incidental findings and added missing (7 total)

NOTE: list was generated using same articles in prompt

List is here:

Head White matter lesions R90.82

Head Arachnoid cyst Q04.3

Head Pituitary microadenoma D35.2

Head Pineal cyst Q04.6

Head Chiari I malformation Q07.0

Neck Thyroid nodule E04.1

Neck Cervical lymphadenopathy R59.0

Neck Parotid gland cyst K11.4

Neck Carotid artery stenosis I65.2

Neck Cervical disc herniation M50.20

Chest Pulmonary nodule R91.1

Chest Mediastinal lymphadenopathy R59.1

Chest Hiatal hernia K44.9

Chest Coronary artery calcification I25.10

Chest Pericardial effusion I31.3

Abdomen Liver cyst K76.89

Abdomen Renal cyst N28.1

Abdomen Adrenal adenoma D35.00

Abdomen Pancreatic cyst K86.2

Abdomen Splenomegaly R16.1

Pelvis Ovarian cyst N83.20

Pelvis Uterine fibroid D25.9

Pelvis Prostatic hypertrophy N40.0

Pelvis Pelvic lymphadenopathy R59.1

Pelvis Bladder diverticulum N32.3

